# Physician-reported practices regarding obesity management in patients aged under 60 years with osteoarthritis in Jordan: a cross-sectional survey

**DOI:** 10.64898/2026.10.27.26366593

**Authors:** Omar M Hanatleh, Abdallah Swaid, Omar F Shatnawi, Nadeem W Bani Hani, Eyad M Bin Tareef, Ali M Ibnian

## Abstract

**Background:** Obesity is an important modifiable risk factor for the development and progression of osteoarthritis (OA), particularly in weight-bearing joints. However, limited information is available regarding physicians’ practices in addressing obesity among patients with OA in Jordan. This study assessed self-reported obesity-management practices among physicians caring for patients aged under 60 years with OA.

**Methods:** We conducted an anonymous, cross-sectional convenience survey between December 2021 and January 2022. Physicians from orthopaedics, family medicine, general internal medicine, and rheumatology were recruited through professional networks and social-media platforms. Participants worked in Ministry of Health, private, university, and military healthcare institutions in Jordan. The questionnaire assessed obesity-related counselling, weight or body mass index (BMI) monitoring, consultation time, and referral for weight-reduction interventions among patients aged under 60 years with obesity and OA.

**Results:** A total of 308 physicians completed the questionnaire. Of these, 162 (52.6%) were orthopedic specialists or trainees and 146 (47.4%) were from other specialties, including primary care, general internal medicine, rheumatology, and general surgery. A total of 138 (44.8%) respondents worked in Ministry of Health healthcare institutions, 87 (28.2%) worked in the private sector, and 83 (26.9%) worked in university or military institutions. Eighty-five respondents (27.6%) reported explaining the effect of obesity on weight-bearing joints to almost all patients with obesity and OA, whereas 149 (48.4%) did so for approximately half of their patients and 74 (24.0%) rarely or never did so. Only 26 respondents (8.4%) reported monitoring weight or BMI in almost all target patients, while 187 (60.7%) reported doing so rarely or in only a few patients. Referral for dietetic or other weight-reduction interventions was reported for almost all target patients by 31 respondents (10.1%), whereas 152 (49.4%) reported rarely or never making such referrals. The remaining respondents reported counselling, monitoring, or referral for approximately half of their target patients.

**Conclusions:** Among participating physicians in Jordan, obesity counselling, weight or BMI monitoring, and referral for weight-reduction interventions were inconsistently incorporated into the management of patients aged under 60 years with OA. These findings support the integration of routine weight assessment, structured obesity counselling, and accessible multidisciplinary referral pathways into OA care. Because this study used convenience sampling and self-reported data, the findings should not be assumed to represent all physicians practising in Jordan.

## 1. Introduction

The worldwide prevalence of obesity continues to rise, contributing to an increasing burden of obesity-related diseases, including osteoarthritis (OA) (1,2). OA is one of the most common joint disorders worldwide and represents a substantial global health burden (3).

Obesity is an important and modifiable risk factor for the development and progression of OA, particularly OA affecting weight-bearing joints such as the knees (4,7). Increased body weight places additional mechanical stress on weight-bearing joints, and knee-joint loading varies according to the type and intensity of physical activity (5). Weight-loss interventions can reduce knee-joint loading and may improve pain and physical function among people with overweight or obesity and knee OA (8,9). Therefore, weight management is an important component of the prevention and treatment of OA.

Physician attitudes and practices related to obesity management have been studied in several clinical contexts, including type 2 diabetes, cardiovascular disease prevention, and primary care (10,13,14,16–18). However, limited evidence is available regarding physicians’ approaches to obesity management among patients with OA, particularly in Jordan.

Jordan has a substantial burden of obesity, and its prevalence has increased over time. In a study by Khader et al., approximately two-thirds of Jordanian adults were classified as overweight or obese (15). Although physicians generally recognise obesity as an important risk factor for several diseases, previous studies have identified gaps in obesity diagnosis, counselling, treatment, and referral practices (16–18). A study from Saudi Arabia also examined primary-care physicians’ attitudes and practices in managing overweight and obesity, providing a relevant regional comparison (19).

In view of the limited evidence regarding obesity management in patients with OA in Jordan, we conducted a cross-sectional survey of physicians practising in Jordan. We assessed self-reported counselling about the relationship between obesity and OA, weight or body mass index monitoring, consultation time, and referral for weight-reduction interventions among patients aged under 60 years with obesity and OA.

Therefore, this study aimed to assess self-reported practices regarding obesity management among physicians caring for patients aged under 60 years with obesity and osteoarthritis in Jordan. Specifically, we evaluated obesity-related counselling, weight or body mass index monitoring, consultation time, and referral for dietetic or other weight-reduction interventions. We also examined whether these practices varied according to physician characteristics, specialty, workplace, and clinical workload.

## 2. Materials and methods

### Study design and participants

We conducted an anonymous, cross-sectional convenience survey of physicians practising in Jordan between December 2021 and January 2022. Eligible participants were consultants and trainees involved in the assessment or management of patients with osteoarthritis. Participants were recruited from orthopaedics, family medicine, general internal medicine, rheumatology, and other relevant specialties working in four healthcare sectors: Ministry of Health, private, university, and military healthcare institutions.

The questionnaire was created using Google Forms and distributed electronically through professional networks and social-media platforms. Participation was voluntary and anonymous.

The questionnaire was developed specifically for this study and was written in English. It included 14 questions divided into two sections. The first section collected demographic and professional information, including sex, age, specialty, workplace, years of clinical experience, and average daily workload. The second section assessed self-reported practices when managing patients aged under 60 years with obesity and osteoarthritis affecting a weight-bearing joint.

The questionnaire assessed whether physicians explained the relationship between obesity and osteoarthritis, measured or monitored body weight or body mass index (BMI), discussed osteoarthritis during the consultation, and referred patients for weight-reduction interventions. Referral options included dietetic assessment and bariatric surgery, where considered clinically appropriate.

### pilot testing of the Study Tool

To assess clarity and face validity, the questionnaire was reviewed by 10 physicians from different specialties and professional backgrounds before distribution. Their feedback was used to improve the wording and clarity of the questions. The questionnaire was developed for this study and was not based on a previously validated instrument.

### Statistical Methodology

All analyses were performed using IBM SPSS Statistics, version 21.0 (IBM Corp., Armonk, NY, USA). Categorical variables are presented as frequencies and percentages. Descriptive statistics were used to summarise participant characteristics and responses to the questionnaire.

Associations between categorical variables were examined using Pearson’s chi-square test or Fisher’s exact test, as appropriate. For ordinal outcomes, the linear-by-linear association test was used where appropriate. Cramér’s V was calculated to describe the strength of associations between nominal categorical variables. All tests were two-sided, and a P-value <0.05 was considered statistically significant. The number of responses included in each analysis was reported, and missing responses were excluded from the relevant analysis.

## 3. Results

### 3.1 Participant characteristics

A total of 308 physicians completed the questionnaire. Of these, 275 (89.3%) were male and 33 (10.7%) were female. One hundred and sixty-two respondents (52.6%) were orthopaedic physicians, while 146 (47.4%) were from primary care, internal medicine, general surgery, or rheumatology. One hundred and fifty-nine respondents (51.6%) had fewer than 8 years of practice and 149 (48.4%) had 8 or more years of practice. One hundred and sixty respondents (51.9%) were aged 33 years or younger and 148 (48.1%) were aged 34 years or older. One hundred and sixty-one respondents (52.3%) reported seeing fewer than 30 patients per day, and 147 (47.7%) reported seeing 30 or more patients per day.

Regarding workplace, 138 respondents (44.8%) worked in Ministry of Health institutions, 87 (28.2%) worked in the private sector, and 83 (26.9%) worked in university or military institutions. Participant characteristics are shown in Table 1.

**Table 1.**
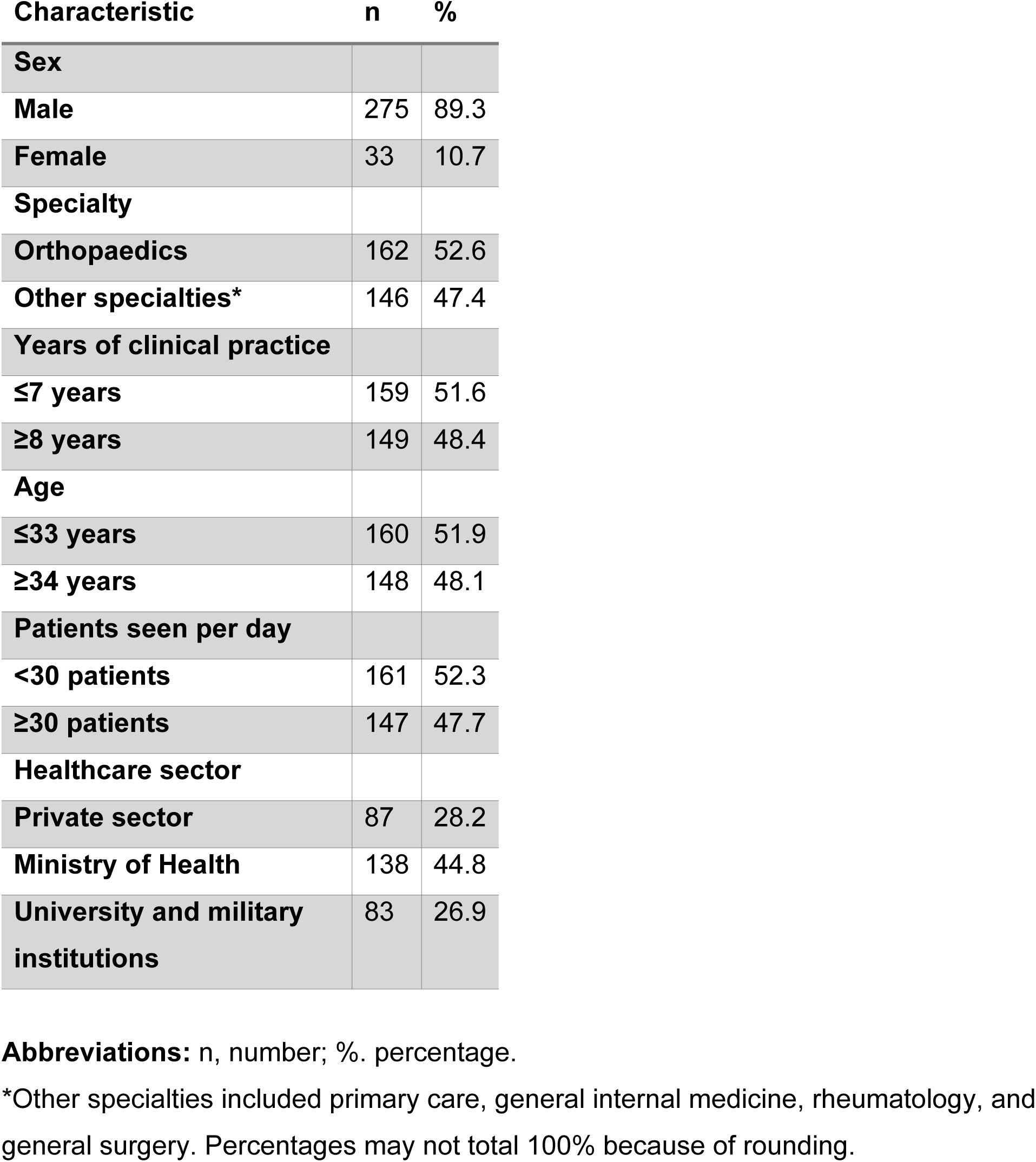
Characteristics of participating physicians (N = 308)

### 3.2 Obesity-related counselling

Eighty-five respondents (27.6%) reported explaining the effect of obesity on weight-bearing-joint OA to almost all patients in the target population (Figure 1). One hundred and forty-nine (48.4%) reported doing so for approximately half of these patients, while 74 (24.0%) reported doing so rarely or in only a few patients.

**Figure 1.**
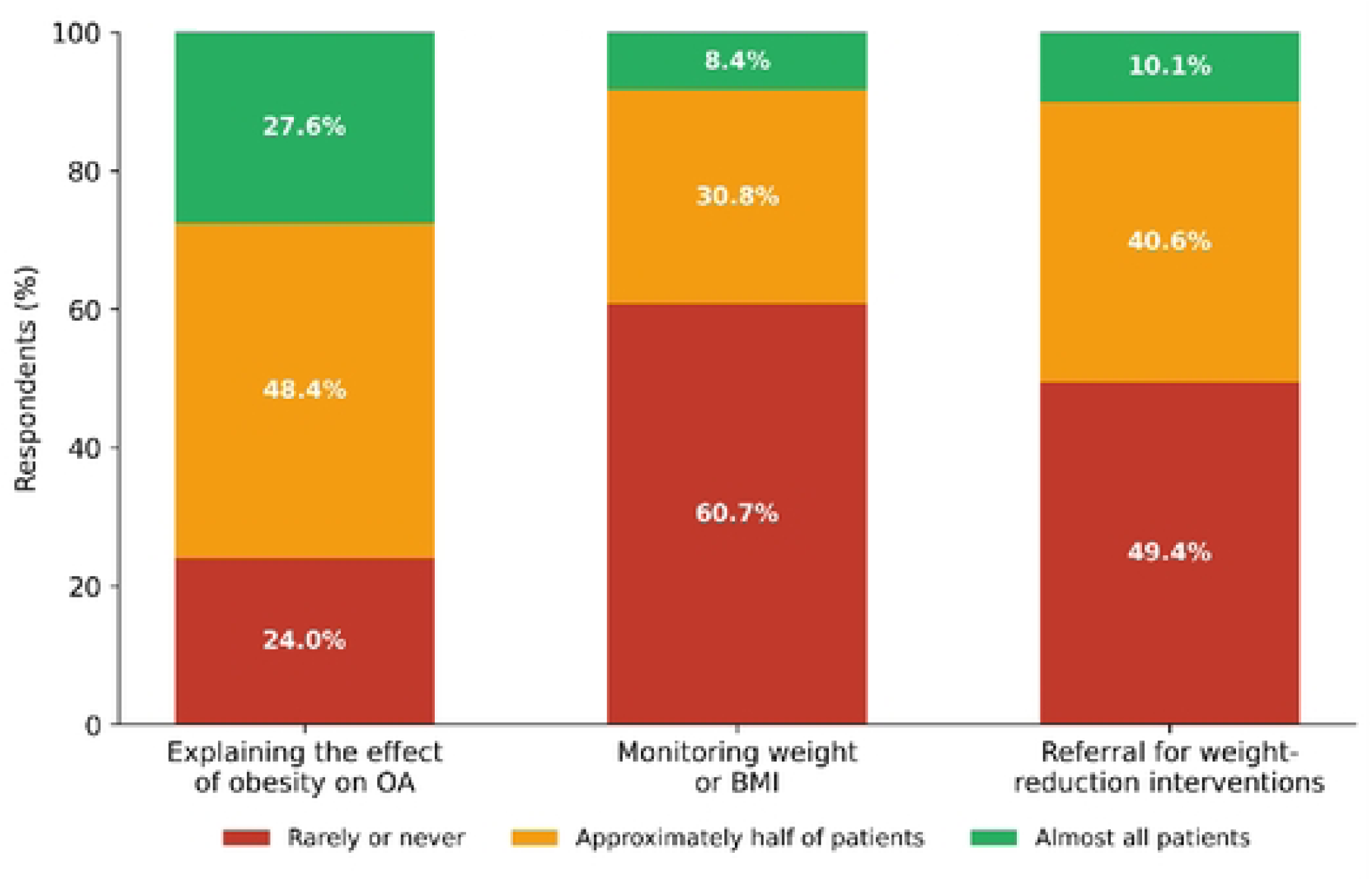
Overall distribution of self-reported obesity-management practices among participating physicians (N = 308). Bars show the proportion of physicians reporting counselling, weight/BMI monitoring, and referral for weight-reduction interventions as “rarely or never,” “approximately half of patients,” or “almost all patients.”

**Figure 2.**
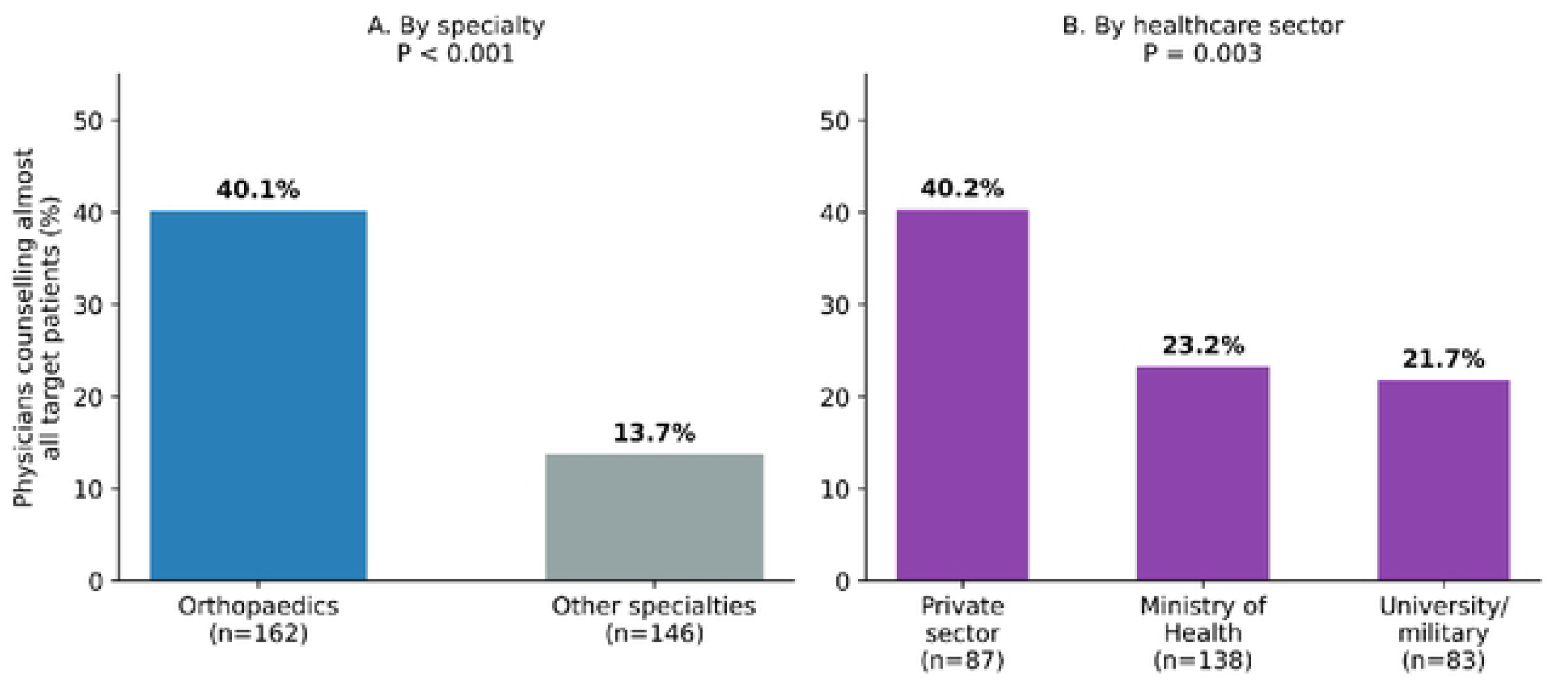
Proportion of physicians reporting obesity-related counselling of almost all target patients, stratified by (A) specialty (orthopaedics vs. other specialties, P < 0.001) and (B) healthcare sector (private, Ministry of Health, university/military, P = 0.003).

Orthopaedic physicians reported counselling almost all target patients more frequently than physicians from other specialties: 65/162 (40.1%) versus 20/146 (13.7%), respectively (P < 0.001). Private-sector physicians also reported counselling almost all target patients more frequently than physicians working in Ministry of Health and other institutions: 35/87 (40.2%), 32/138 (23.2%), and 18/83 (21.7%), respectively (P = 0.003) (Figure).

Male physicians, physicians aged 34 years or older, and physicians with 8 or more years of practice also reported counselling almost all target patients more frequently than their respective comparison groups: 81/275 (29.5%) versus 4/33 (12.1%) (P = 0.030); 49/148 (33.1%) versus 36/160 (22.5%) (P = 0.005); and 49/149 (32.9%) versus 36/159 (22.6%) (P = 0.013), respectively. These are unadjusted associations and should not be interpreted as independent effects.

### 3.3 Weight and BMI monitoring

Twenty-six respondents (8.4%) reported monitoring weight or BMI in almost all target patients. Ninety-five (30.8%) reported doing so for approximately half of target patients, while 187 (60.7%) reported doing so rarely or in only a few patients.

Orthopaedic physicians reported monitoring weight or BMI in almost all target patients less frequently than physicians from other specialties: 7/162 (4.3%) versus 19/146 (13.0%), respectively (P = 0.020). No, statistically significant associations were reported for sex, age, years of practice, workload, or workplace in the supplied analysis. The corresponding exact counts and denominators in Table 2 and Table 3.

**Table 2.**
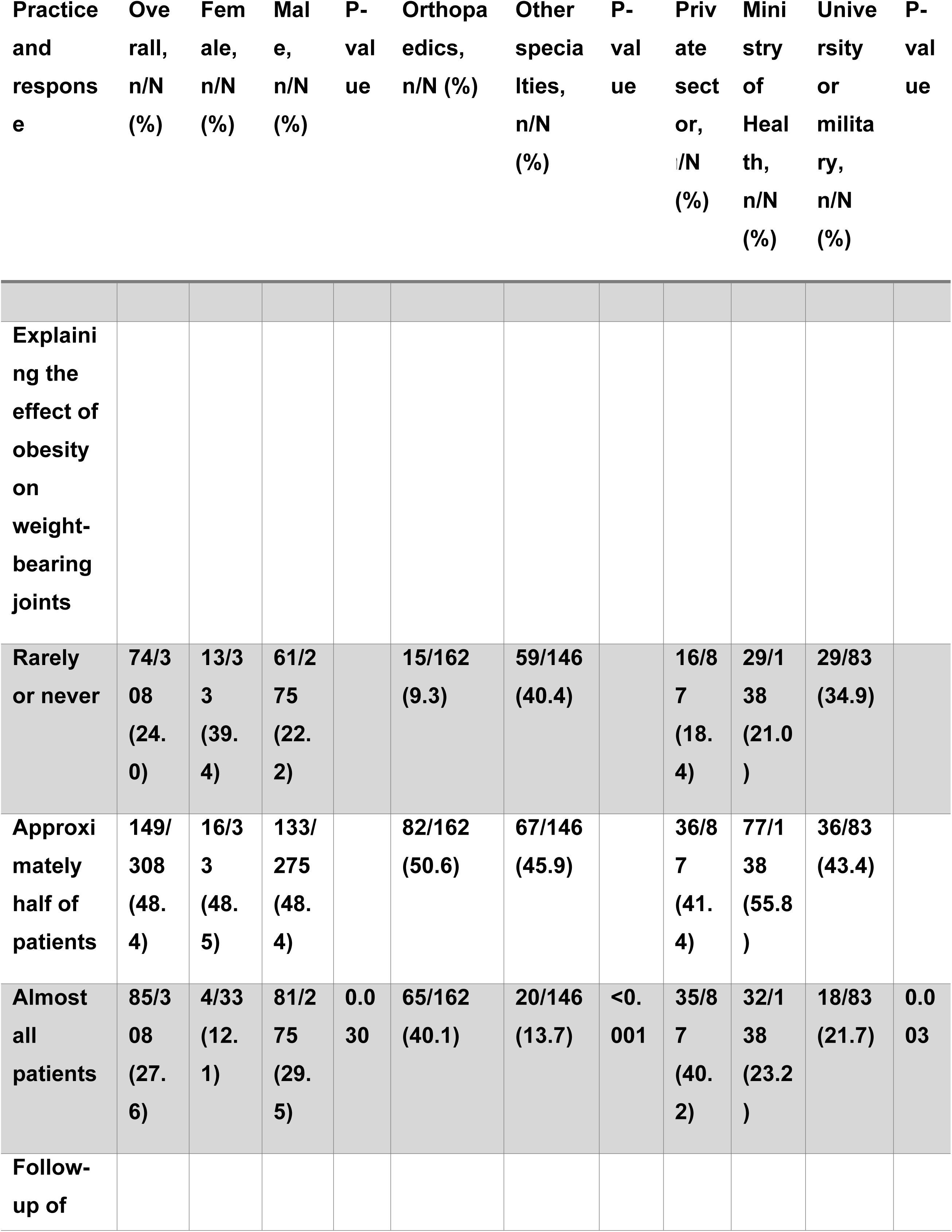
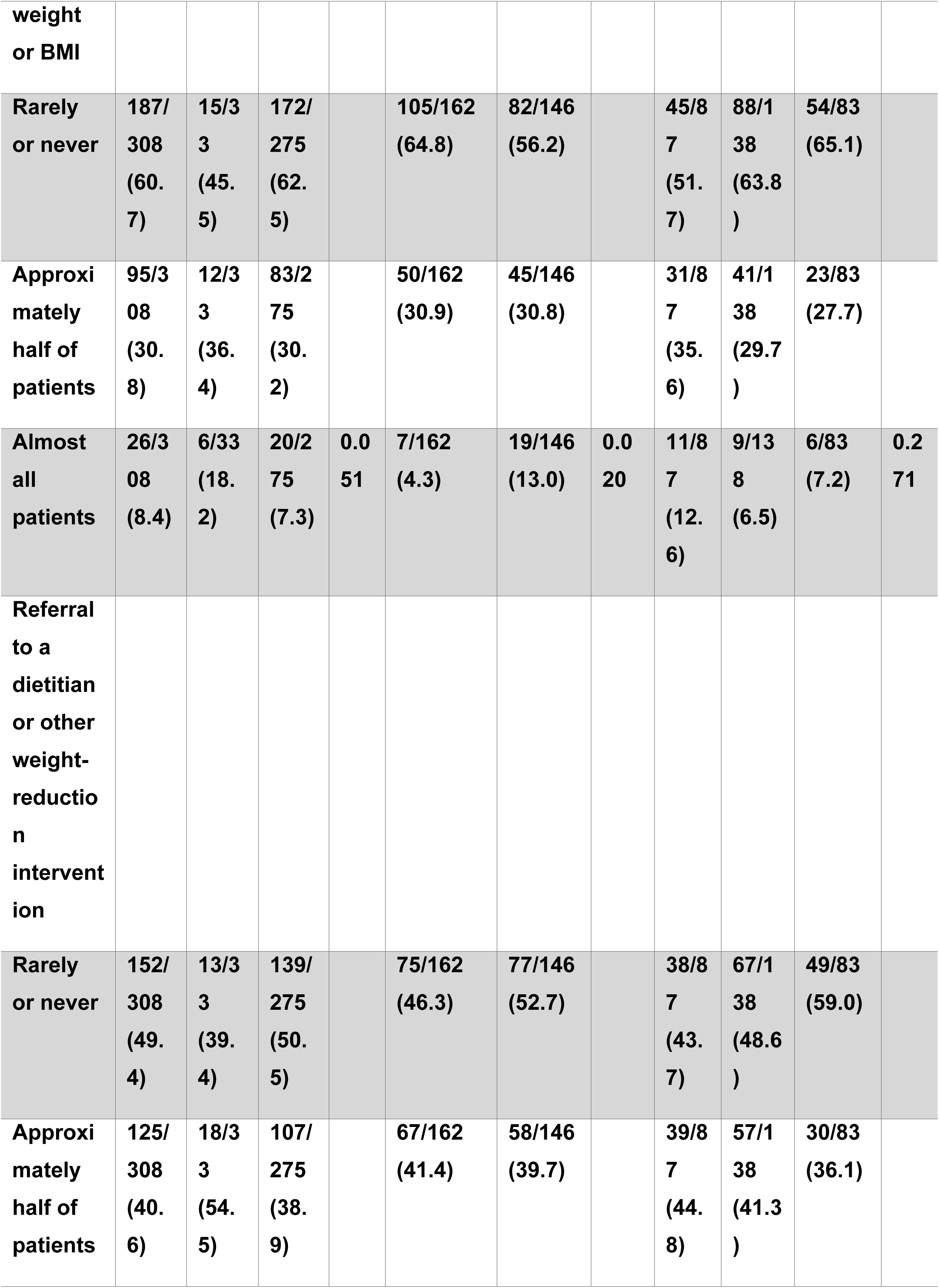
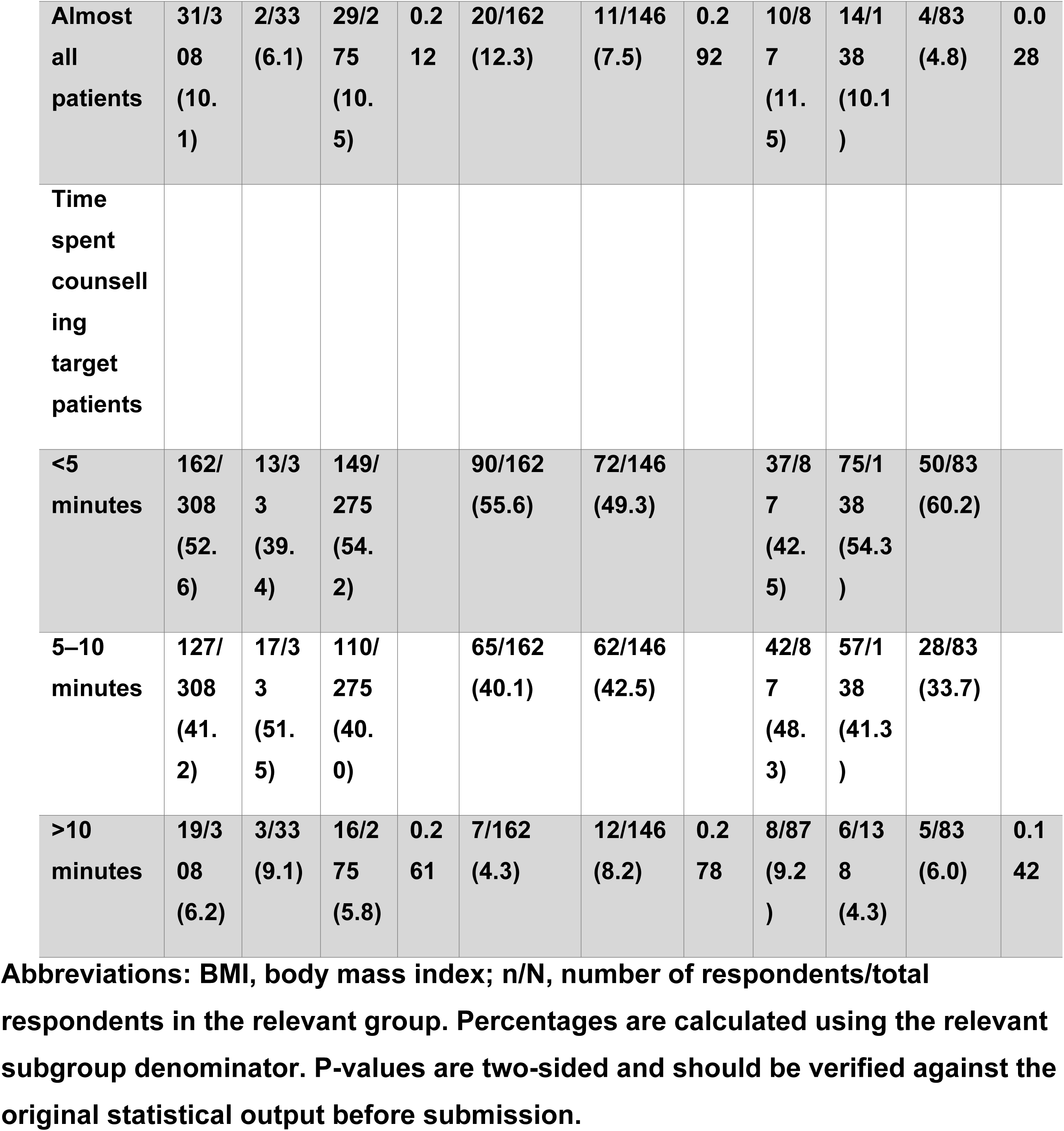
Associations of sex, specialty, and workplace with selected obesity-management practices.

**Table 3:**
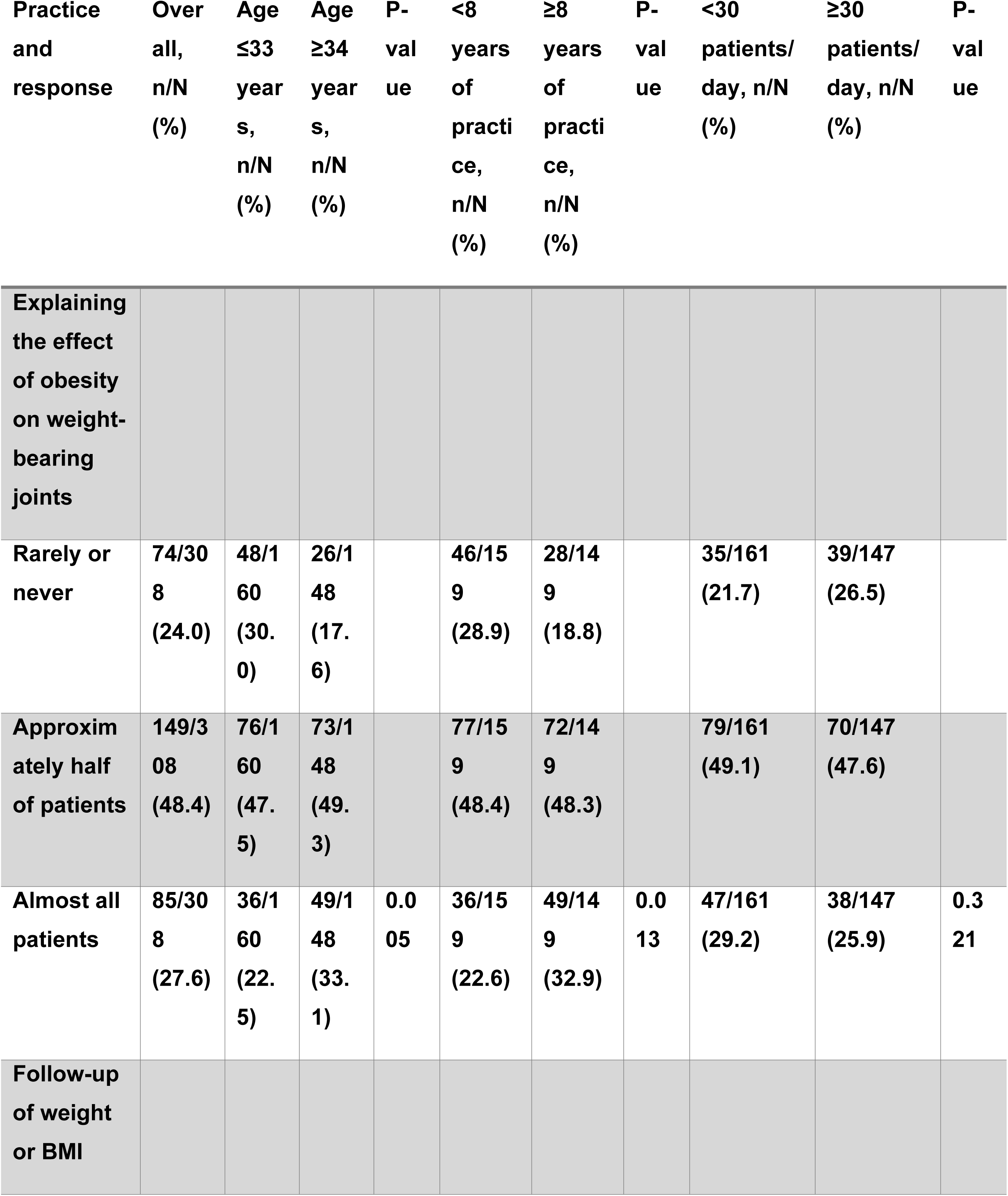
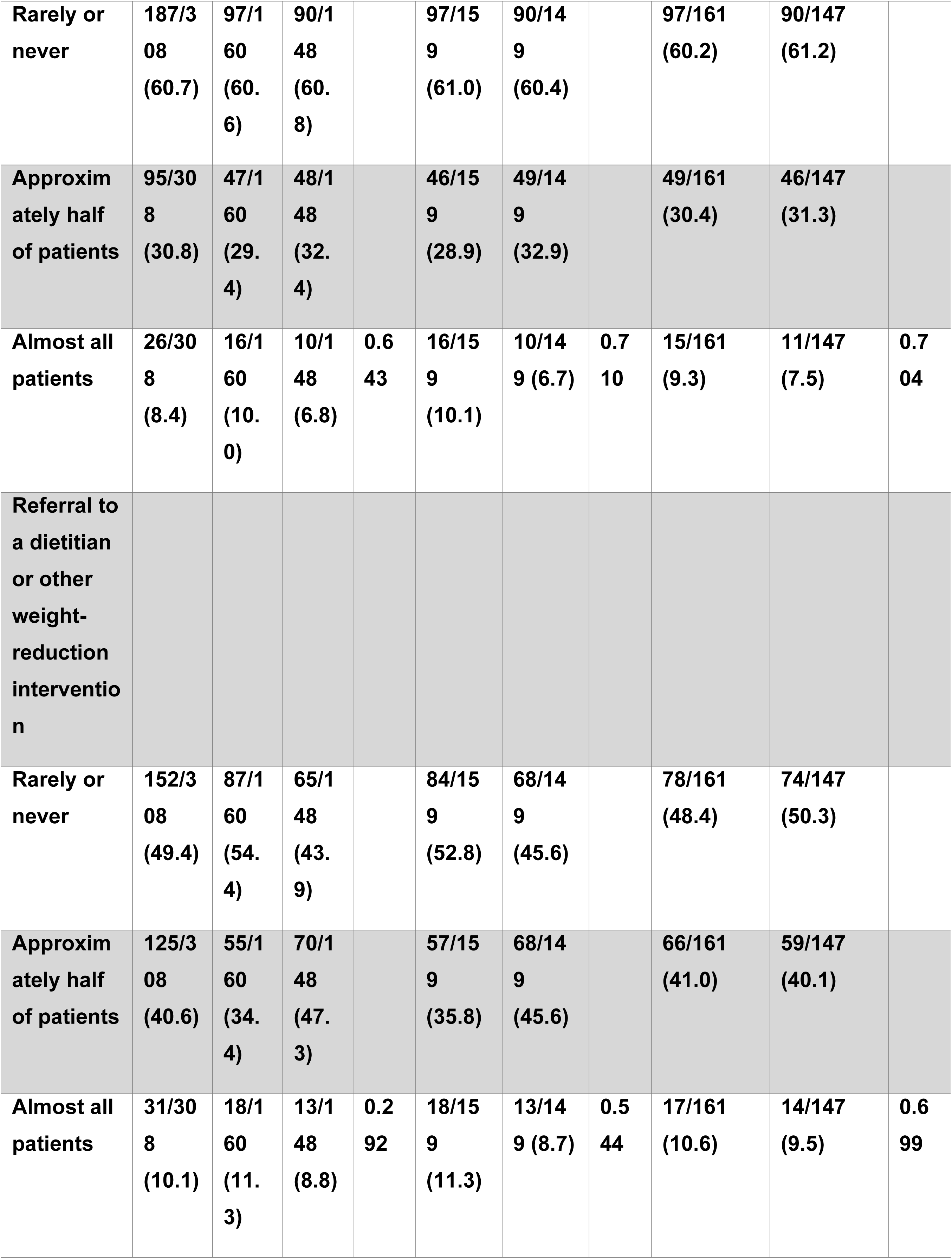
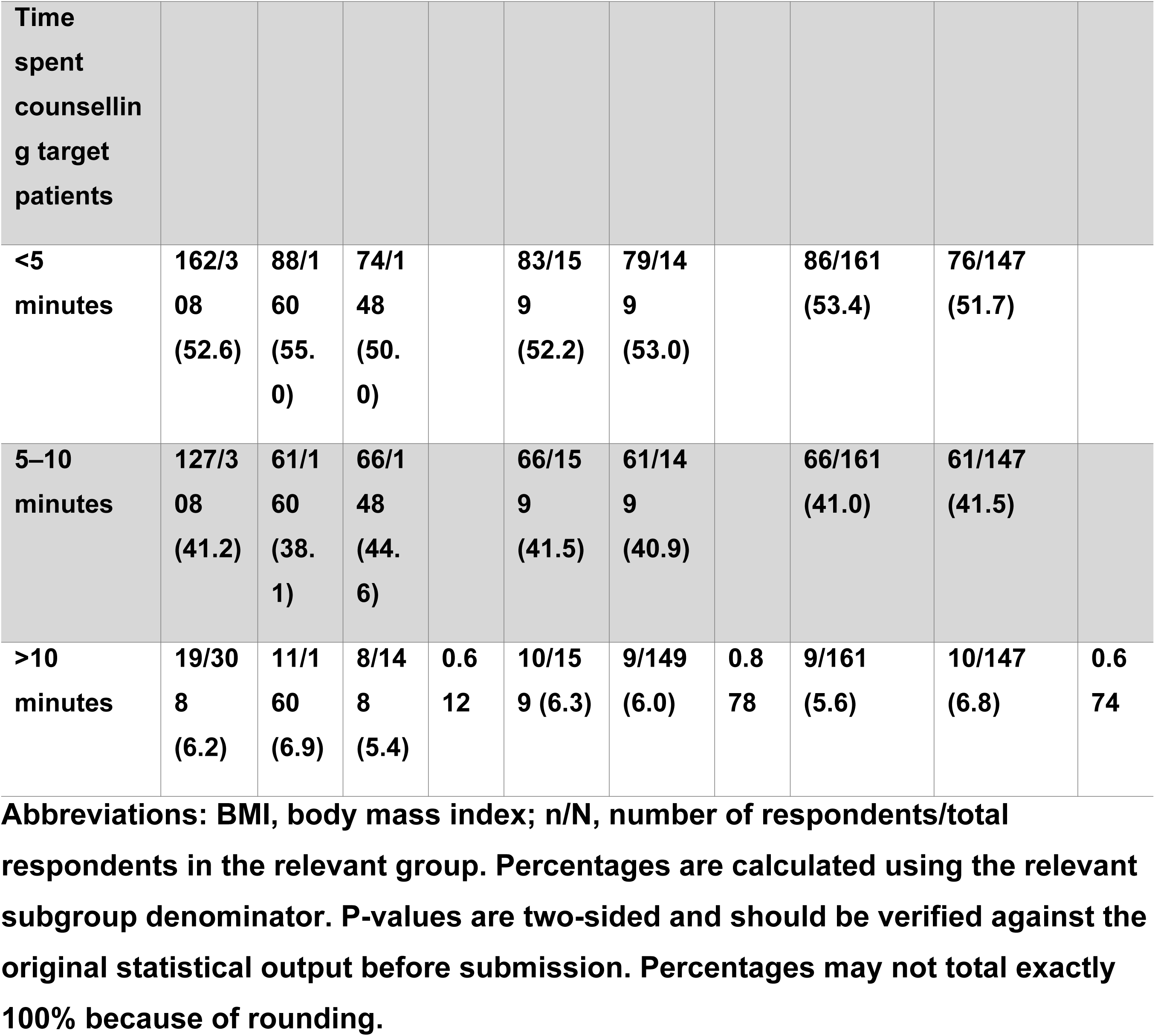
The association between physician’s age, experience, and working load, and working place on their attitude in management of obese young patients with osteoarthritis (the target patients).

### 3.4 Referral for weight-reduction interventions

Thirty-one respondents (10.1%) reported referring almost all target patients to a dietitian or for another weight-reduction intervention. One hundred and twenty-five (40.6%) reported doing so for approximately half of target patients, while 152 (49.4%) reported doing so rarely or in only a few patients.

Referral patterns differed by workplace (P = 0.028). The proportion reporting referral of almost all target patients was 10/87 (11.5%) among private-sector physicians, 14/138 (10.1%) among Ministry of Health physicians, and 4/83 (4.8%) among physicians working in university or military institutions presented in Tables 2 and 3.

### 3.5 Consultation time

One hundred and sixty-two respondents (52.8%) reported spending fewer than 5 minutes counselling target patients, 127 (41.2%) reported spending 5–10 minutes, and 19 (6.2%) reported spending more than 10 minutes (Tables 2 and 3).

## 4. Discussion

This cross-sectional survey found that obesity-related counselling, weight monitoring, and referral for weight-reduction interventions were inconsistently reported by participating physicians caring for patients aged under 60 years with obesity and weight-bearing-joint osteoarthritis in Jordan. Only approximately one-quarter of respondents reported explaining the importance of obesity management to almost all target patients, fewer than one in ten reported monitoring weight or BMI in almost all target patients, and approximately one in ten reported referring almost all target patients for dietetic or other weight-reduction interventions.

Orthopaedic physicians reported counselling almost all target patients more frequently than physicians from other specialties. This finding may reflect their frequent involvement in the care of patients with weight-bearing-joint osteoarthritis. However, because the study was cross-sectional and the analyses were unadjusted, these findings should be interpreted as associations rather than independent effects. The observed differences between healthcare sectors may reflect variation in access to dietitians, consultation structure, referral pathways, institutional policies, workload, or patient characteristics.

A previous study from Saudi Arabia examined primary-care physicians’ general practices in managing overweight and obesity and reported that many physicians counselled patients about diet and exercise, although fewer considered themselves competent in obesity management (19). The Saudi study differed from the present study because it addressed obesity management more broadly, did not focus specifically on patients with osteoarthritis, and did not include a predominantly orthopaedic sample.

Similarly, a study conducted in two public hospitals in the United States reported that weight-loss counselling was documented for many patients, whereas specific weight-loss recommendations were provided less frequently (20). Although these studies used different populations and methods, their findings and ours suggest a possible gap between discussing obesity and implementing specific weight-management interventions.

The reasons for this gap are likely to be multifactorial. Counselling may be difficult to translate into treatment when physicians have limited consultation time, inadequate training, restricted access to dietitians or structured weight-management programmes, or unclear referral pathways. Patient acceptance, concerns about weight stigma, healthcare costs, and the perceived responsibility of different specialties may also influence whether obesity is addressed. These potential explanations were not directly assessed in the present survey and should be examined in future research.

### 4.1 Clinical implications

The findings support several practical measures for strengthening obesity management within osteoarthritis care:

routine recording of weight and BMI during the initial osteoarthritis assessment;
periodic weight monitoring during follow-up;
brief, non-stigmatising counselling about the relationship between obesity and osteoarthritis;
clear referral pathways to dietitians and multidisciplinary weight-management services;
and
collaboration between orthopaedic, primary-care, rheumatology, internal medicine, and obesity-management services.

These recommendations should be adapted to local service availability and should form part of shared decision-making with patients. This study did not assess whether referral services were accessible, whether patients accepted referrals, or whether counselling resulted in weight loss, symptom improvement, or slower osteoarthritis progression.

### 4.2 Strengths and limitations

A strength of this study is that it examined selected obesity-management practices in a clinically relevant population and included physicians from several specialties and healthcare sectors in Jordan.

Several limitations should be considered. First, participants were recruited through a convenience-based electronic strategy using professional networks and social-media platforms. A complete sampling frame and denominator of eligible physicians were not available, and a formal response rate could not be calculated. Selection and non-response bias are therefore possible, and the findings should not be assumed to represent all physicians practising in Jordan. Second, the outcomes were self-reported and may be affected by recall and social-desirability bias. Third, the questionnaire was developed for this study and was not based on a previously validated attitude or practice scale. Fourth, the study assessed selected reported practices rather than comprehensive obesity management and did not evaluate patient outcomes, actual weight loss, osteoarthritis progression, or access to referral services.

## Data Availability

will be uploaded as supplementary

## 5. Conclusion

Obesity-related counselling, weight monitoring, and referral for weight-reduction interventions were inconsistently reported by physicians caring for patients aged under 60 years with obesity and weight-bearing-joint OA in Jordan. Routine BMI assessment, structured counselling, and accessible referral pathways may strengthen obesity management in this population. Larger studies using representative sampling, validated instruments, multivariable analysis, and objective measures of clinical practice are warranted.

## Declaration of consent

The authors declare that they have obtained consent from all physicians included in the study.

acknowledgments: None

## Data Availability Statement

The data that supports the findings of this study are available from the corresponding author upon reasonable request

This study did not receive any specific grant from funding agencies in the public, commercial, or non-profit sectors

“All authors gave final approval of the version to be published; have agreed on the journal to which the article has been submitted; and agree to be accountable for all aspects of the work.“

Artificial intelligence (AI) was used to correct grammar and punctuation and to improve the flow of the writing. AI was not used to develop the study methodology, perform the analysis, or write the initial manuscript. All AI-recommended changes were subsequently reviewed and approved by all authors.

## Authors Contributions

**OH:** Was responsible for designing the study questionnaire, reviewing the data, and preparing the data for statistical analysis. He contributed to writing the draft of the manuscript. He also revised and approved the final manuscript.

**AS:** Contributed to reviewing and critically revising the manuscript, checking the presentation and clarity of the content, coordinating the final manuscript preparation.

**OS:** Was responsible for designing the study questionnaire, reviewing the data, and preparing the data for statistical analysis. He contributed to writing the draft of the manuscript. He also revised and approved the final manuscript.

**NB:** Was responsible for writing the draft of the manuscript. He contributed to reviewing the final manuscript and preparing the manuscript format for publication.

**EB:** Was responsible for designing and checking the study questionnaire. He contributed to the final check and proofreading of the manuscript. He also revised the manuscript and approved the final version.

**AI:** Was responsible for designing the study protocol, designing the study questionnaire, writing the manuscript, reviewing the data, preparing the data for analysis, extracting and analysing data, writing the draft manuscript, proofreading the final version of the manuscript, and preparing the manuscript format for publication.

## References

1. Chooi YC, Ding C, Magkos F. The epidemiology of obesity. Metabolism. 2019 Mar 1;92:6–10.

2. Binvignat M, Sellam J, Berenbaum F, Felson DT. The role of obesity and adipose tissue dysfunction in osteoarthritis pain. Nat Rev Rheumatol 2024 209 [Internet]. 2024 Aug 7 [cited 2026 Aug 13];20(9):565–84. Available from: https://www.nature.com/articles/s41584-024-01143-3

3. Safiri S, Kolahi AA, Smith E, Hill C, Bettampadi D, Mansournia MA, et al. Global, regional and national burden of osteoarthritis 1990-2017: a systematic analysis of the Global Burden of Disease Study 2017. Ann Rheum Dis [Internet]. 2020 [cited 2021 Dec 5];79(6). Available from: https://pubmed-ncbi-nlm-nih-gov.go.libproxy.wakehealth.edu/32398285/

4. Lespasio MJ, Piuzzi NS, Husni ME, Muschler GF, Guarino A, Mont MA. Knee Osteoarthritis: A Primer. Perm J [Internet]. 2017 [cited 2021 Dec 11];21. Available from: https://pubmed-ncbi-nlm-nih-gov.go.libproxy.wakehealth.edu/29035179/

5. Kuster MS, Wood GA, Stachowiak GW, Gächter A. Joint load considerations in total knee replacement. J Bone Joint Surg Br [Internet]. 1997 Jan [cited 2022 Feb 11];79(1):109–13. Available from: https://pubmed.ncbi.nlm.nih.gov/9020457/

6. What you need to know about osteoarthritis | Ottobock US [Internet]. [cited 2022 Feb 11]. Available from: https://www.ottobockus.com/orthotics/osteoarthritis-less-pain.-more-life./what-you-need-to-know-about-osteoarthritis.html

7. King LK, March L, Anandacoomarasamy A. Obesity & osteoarthritis. Indian J Med Res [Internet]. 2013 Aug [cited 2022 Oct 15];138(2):185. Available from: /pmc/articles/PMC3788203/

8. Messier SP, Gutekunst DJ, Davis C, DeVita P. Weight loss reduces knee-joint loads in overweight and obese older adults with knee osteoarthritis. Arthritis Rheum [Internet]. 2005 Jul [cited 2022 Mar 20];52(7):2026–32. Available from: https://pubmed.ncbi.nlm.nih.gov/15986358/

9. Shahid A, Thirumaran AJ, Christensen R, Venkatesha V, Henriksen M, Bowden JL, et al. Comparison of weight loss interventions in overweight and obese adults with knee osteoarthritis: A systematic review and network meta-analysis of randomized trials. Osteoarthr Cartil [Internet]. 2025 Apr 1 [cited 2026 Aug 13];33(4):518–29. Available from: https://www.oarsijournal.com/action/showFullText?pii=S1063458424013670

10. Sarwer DB, Ritter S, Wadden TA, Spitzer JC, Vetter ML, Moore RH. Physicians’ Attitudes about Referring their Type 2 Diabetes Patients for Bariatric Surgery. Surg Obes Relat Dis [Internet]. 2012 Jul [cited 2022 Mar 20];8(4):381–6. Available from: /pmc/articles/PMC3865449/

11. Bramlage P, Pittrow D, Wittchen HU, Kirch W, Boehler S, Lehnert H, et al. Hypertension in overweight and obese primary care patients is highly prevalent and poorly controlled. Am J Hypertens [Internet]. 2004 Oct 1 [cited 2022 Mar 20];17(10):904–10. Available from: https://academic.oup.com/ajh/article/17/10/904/106819

12. Engin A. The Definition and Prevalence of Obesity and Metabolic Syndrome. Adv Exp Med Biol [Internet]. 2017 [cited 2021 Dec 4];960:1–17. Available from: https://pubmed-ncbi-nlm-nih-gov.go.libproxy.wakehealth.edu/28585193/

13. Hobbs FDR, Erhardt L. Acceptance of guideline recommendations and perceived implementation of coronary heart disease prevention among primary care physicians in five European countries: the Reassessing European Attitudes about Cardiovascular Treatment (REACT) survey. Fam Pract [Internet]. 2002 Dec 1 [cited 2022 Mar 23];19(6):596–604. Available from: https://academic.oup.com/fampra/article/19/6/596/477706

14. Hølund U, Thomassen A, Boysen G, Charles P, Eriksen EF, Overvad K, et al. Importance of diet and sex in prevention of coronary artery disease, cancer, osteoporosis, and overweight or underweight: a study of attitudes and practices of Danish primary care physicians. Am J Clin Nutr [Internet]. 1997 Jun 1 [cited 2022 Mar 23];65(6):2004S–2006S. Available from: https://academic.oup.com/ajcn/article/65/6/2004S/4655666

15. Khader Y, Batieha A, Ajlouni H, El-Khateeb M, Ajlouni K. Obesity in Jordan: prevalence, associated factors, comorbidities, and change in prevalence over ten years. Metab Syndr Relat Disord [Internet]. 2008 Jun 1 [cited 2022 Feb 5];6(2):113–20. Available from: https://pubmed.ncbi.nlm.nih.gov/18510436/

16. Bleich SN, Pickett-Blakely O, Cooper LA. Physician practice patterns of obesity diagnosis and weight-related counseling. Patient Educ Couns. 2011 Jan 1;82(1):123–9.

17. Kristeller JL, Hoerr RA. Physician Attitudes toward Managing Obesity: Differences among Six Specialty Groups. Prev Med (Baltim). 1997 Jul 1;26(4):542–9.

18. Brandsma LL. Physician and Patient Attitudes Toward Obesity. 10.1080/10640260590919134 [Internet]. 2006 Mar [cited 2022 Apr 1];13(2):201–11. Available from: https://www.tandfonline.com/doi/abs/10.1080/10640260590919134

19. Fadhel Y, Alshammari F, Yf A-S. Attitudes and Practices of Primary Care Physicians in the Management of Overweight and Obesity in Eastern Saudi Arabia. Int J Health Sci (Qassim) [Internet]. 2014 Apr [cited 2022 Mar 6];8(2):151. Available from: /pmc/articles/PMC4166987/

20. Physicians’ Weight Loss Counseling in Two Public Hospital Pr… : Academic Medicine [Internet]. [cited 2022 Mar 6]. Available from: https://journals.lww.com/academicmedicine/Fulltext/2004/02000/Physicians_Weight_Loss_Counseling_in_Two_Public.12.aspx

